# A Randomized Non-Inferiority Trial of an eHealth Delivery Alternative for Cancer Genetic Testing for Hereditary Cancer (eREACH2)

**DOI:** 10.64898/2026.09.01.26361920

**Authors:** Kimberley T. Lee, Brian Egleston, Dominique Fetzer, Susan M. Domchek, Linda Fleisher, Kuang-Yi Wen, Lynne Wagner, J. Scott Roberts, Sarah Howe, Cara Cacioppo, Janice Christiansen, Kelsey Karpink, Enida Selmani, Evelyn Mastaglio, Michelle Weinberg, Elisabeth Wood, Justin Feng, Samantha John, K Schweickert, B McLeod, Angela R. Bradbury

**Author notes:** **Research support:** National Cancer Institute U01CA243702 (no role in design, conduct, analysis, or reporting of trial). **Funding:** National Cancer Institute U01CA243702. **Trial registration:** This protocol was registered at clinicaltrials.gov (NCT05427240) on 6/7/2022.

## Abstract

**Background:** Many at-risk patients lack access to genetic services due to a genetic counselor (GC) workforce shortage. Little is known about how digital alternatives impact patients with and without cancer who meet criteria for genetic testing.

**Methods:** eREACH2 is a randomized 4-arm non-inferiority trial where pre-test (visit 1) and/or return of results (visit 2) GC counseling was replaced with a patient-centered digital intervention. Arms include: A (GC/GC), B (GC/digital), C (digital/GC) and D (digital/digital). Primary outcomes were non-inferiority in uptake of genetic services and change in genetic knowledge and general anxiety from baseline to post-disclosure of results (T0-T2). Secondary cognitive and affective outcomes were assessed using non-inferiority ANOVAs and equivalency chi-squared tests in intention-to-treat and per-protocol analyses.

**Findings:** 773 participants were recruited nationwide; 46.6% from rural areas. Mean age was 51 years (range 20-87), 13% male, 12% non-white, 29% had less than a college education, and 33% had a personal history of cancer. 584 (76%) patients completed testing (14% had a positive result, 16% had a VUS). In the primary ITT analyses, we met the non-inferiority for uptake of genetic services and anxiety, but results were inconclusive for knowledge. Secondary outcomes were heterogeneous across arms. Arm C demonstrated consistently favorable effects, while Arms B and D showed less favorable outcomes in select domains (e.g. satisfaction and MICRA). Patients who received positive or VUS results via digital disclosure had significantly higher MICRA scores — indicating greater negative response to testing.

**Interpretation:** In this large, randomized trial of patients with and without cancer, the eREACH intervention was effective for pre-test counseling, but inconclusive for digital disclosure of results. Exploratory analyses suggest that digital delivery could be a reasonable alternative for individuals receiving negative results, while those receiving positive or VUS results may derive some short-term psychosocial benefit from GC disclosure.

## INTRODUCTION

Germline cancer genetic testing has important implications for primary and secondary cancer prevention for persons with pathogenic variants (PV).[1, 2] Germline cancer genetic testing also has the potential to personalize cancer treatment with the approval of treatments such as poly adenosine diphosphate ribose polymerase (PARP) inhibitors for patients with a germline *BRCA1/2* PV or likely PV and breast, ovarian, prostate, and pancreatic cancers.[3, 4] Yet, many at-risk patients do not have access to genetic services, leaving many genetic carriers unidentified.[5–7] Access to genetic counselors (GC) is limited in many areas in the US, and the traditional delivery model of pre- and post-test counseling with a GC will not support the rising indications for germline cancer genetic testing.[8, 9] Thus, there is a pressing need to evaluate alternative delivery models to increase access to and efficiency of germline testing for therapeutic indications.

Several studies have identified favorable outcomes with remote provision of genetic counseling services either by phone or videoconference.[10–14] While remote telegenetic services can increase access to services, this model alone does not address the growing indications for clinical genetic testing and the limited genetic counselor (GC) workforce. The recent eREACH study found that digital delivery of pre-test counseling or post-test disclosure of results was non-inferior to two visits with a GC among patients with certain metastatic cancers.[15] Thus, interactive, patient-centered digital interventions, used in collaboration with genetic specialists may be an alternative way of addressing the workforce shortage while maintaining adequate patient outcomes.[10, 15–18]

This study, a **R**andomized Hybrid Type I Effectiveness-Implementation Study of an **e**Health Delivery **A**lternative for **C**ancer Genetic Testing for **H**ereditary Cancer (eREACH2) is a 2×2 non-inferiority study randomizing real-world clinical patients who meet National Comprehensive Cancer Network (NCCN) or American Society of Clinical Oncology (ASCO) guidelines for germline genetic testing to a patient-centered multimodal digital intervention versus traditional pre-test (visit 1: education and consent) and/or post-test (visit 2: disclosure) counseling delivered by a GC. The primary objective was to assess whether the digital intervention would result in equal or improved uptake of genetic services and cognitive and affective outcomes (knowledge and anxiety) from baseline to post-disclosure assessment.

## MATERIALS AND METHODS

### Participants

Participants were English speaking adults who were 18 years of age or older, had no prior clinical germline genetic testing, and met current NCCN or ASCO guidelines for germline genetic testing. Participants were recruited from Penn Medicine clinics, regional community oncology clinics affiliated with the Penn Telegenetics program, and social media ads through Penn and advocacy organizations (breastcancer.org). Patients with prior germline genetic testing, uncorrected or uncompensated speech defects, uncontrolled psychiatric/mental condition or cognitive deficits rendering the individual unable to understand study goals or tasks were excluded. The study protocol was approved by the University of Pennsylvania’s institutional review board.

### Digital Interventions for Pre-test education and disclosure of results

As previously described, the eREACH digital interventions are theoretically and GC informed, user-tested, interactive patient-centered digital interventions for germline genetic testing.[19] Briefly, the digital intervention includes Tier 1, indispensable information presented to all users, and optional Tier 2 content (more in-depth information, examples and/or videos). The pre-test intervention included eight modules and participants were asked to record their testing decision. The digital result disclosure (visit 2) intervention includes four modules as previously described. Participants assigned to the digital intervention could request a GC visit at any time.

### Randomization and Procedures

After informed consent and the baseline survey (T0), participants were randomized to one of the four arms, stratified by gender and cancer type and using a permuted block design.

In the intervention arms (Arms B-D) the traditional standard-of-care pre-test (visit 1) and/or return of results (visit 2) counseling delivered by a GC was replaced with the digital intervention to give Arms A (GC/GC), B (GC/digital), C (digital/GC) and D (digital/digital). Participants assigned to a digital visit could request a visit with a GC if preferred. All visits with a GC were conducted by phone or videoconference. Full study procedures have been previously described.[20]

All genetic testing was sent to a commercial laboratory and covered by insurance. Participants could choose a smaller panel based on personal and family history and focused on genes that have strong evidence to change cancer care or screening, a larger or more comprehensive panel which could include genes of unclear immediate value but that could have a potential impact on cancer treatment or screening in the future, or a customized panel based on other health concerns or a desire to exclude particular genes.

### Primary Outcomes

Theoretically informed outcomes to evaluate cognitive, affective and behavioral outcomes of digital alternatives as compared to meeting with a GC were collected at baseline (T0), after Visit 1 (T1) and Visit 2 (T2), at 6 months (T3), and at 12 months (T4). Participants received a $10 gift card for each survey completed.

The primary outcomes included:

*1)* Uptake of genetic services.
*2) Knowledge of genetic disease* evaluated with the 16-item The KnowGene Scale (range 0-16, with 2 points representing a clinically significant difference) (alpha=0.80-0.83).[21]
*3)* Anxiety assessed by the 4-item Patient Reported Outcomes Measurement Information System (PROMIS) Anxiety measure (alpha=0.89-0.93, with 3.5 difference in t-score being clinically relevant).[22]

### Secondary Outcomes

Secondary outcomes have been previously described [19] and included recall of genetic test results and PROs 1) Depression (4-item PROMIS Depression, alpha=0.89-0.93),[22] 2) Disease-specific distress (Impact of Events Scale, range 0-40, alpha=0.90-0.92),[23, 24] 3) Satisfaction with genetic services (range 8-40, alpha=0.83),[10, 15, 25] 4) Responses to testing (Multi-dimensional Impact of Cancer Risk Assessment Questionnaire, range 0-95, alpha 0.75-0.83),[26] and 5) Decisional regret (5-item Decision Regret Scale, range 5-25, alpha 0.77-0.94).[27] GCs recorded time spent on patient care activities for a subset of participants to estimate provider time by arm.

Moderators were collected at baseline and included attitudes to genetic testing, race/ethnicity, education, marital status, gender and age. The Social Vulnerability Index (SVI) is a place-based index designed to quantify communities experiencing social vulnerability based on socioeconomic status, household characteristics, racial and ethnic minority status, and housing type and transportation and was identified using participant zip code.[28] Higher scores indicate greater social vulnerability. Health literacy was assessed with the Brief Health Literacy Screen.[29] Family history of cancer was defined as cancers of the breast, ovary, pancreas, or colorectum in a first or second-degree relative.

### Statistical Analysis

The primary outcomes included changes in knowledge and anxiety from the baseline to the post-disclosure period (T0-T2) and uptake of genetic services. Secondary outcomes included depression, cancer specific distress, uncertainty, change in treatment plan, communication of results and provider time. In order to maximize power in a four-am study, we used a joint test that the three eHealth arms were noninferior to Arm A (GC/GC), as described in our supplementary material.[30] Our design had 88% noninferiority power and 1.67% Type I error to account for three primary endpoints. For knowledge and anxiety, we assumed null hypotheses of -0.20 standardized worsening in Arms B and C and -0.39 standardized worsening in arm D. For uptake, we assumed a null of 9% worse uptake in arms B and C and 19% worse uptake in arm D. Our alternative hypotheses assumed no differences among arms. To rule out heterogeneous qualitative effects among the four arms, such as the intervention being beneficial in some arms, but harmful in others, we examined pairwise comparisons among the arms using T-tests and chi-squared tests in secondary analyses. We used linear and logistic regressions to assess moderators of intervention effects. We used multiple imputation for the outcome analyses.[31]

## RESULTS

### Study Participants

Enrollment, randomization, and survey completion are shown in **Figure 1**. Of 853 participants who consented to the study, 785 (92%) completed the baseline survey, and 773 (90.6%) were randomized. The mean age of the participants was 51 years, 88.0% were white, 4.0% were Black, 6.1% were Hispanic/Latinx, and 29.2% had less than a college degree. Participants were recruited from 46 states across the United States, 46.6% were from rural areas, 48.9% were referred from a community site and 36.1% were recruited from social media or otherwise self-referred. Thirty-two percent of participants had a personal history of cancer and there was a statistically significant difference in personal history of cancer between randomized groups, and 115 (14.9%) had a known familial mutation (**Table 1**). Among those tested, most participants (69.4%) had a negative result, 91 (15.6%) had a positive result and 88 (15.0%) had a VUS.

**Figure 1.**
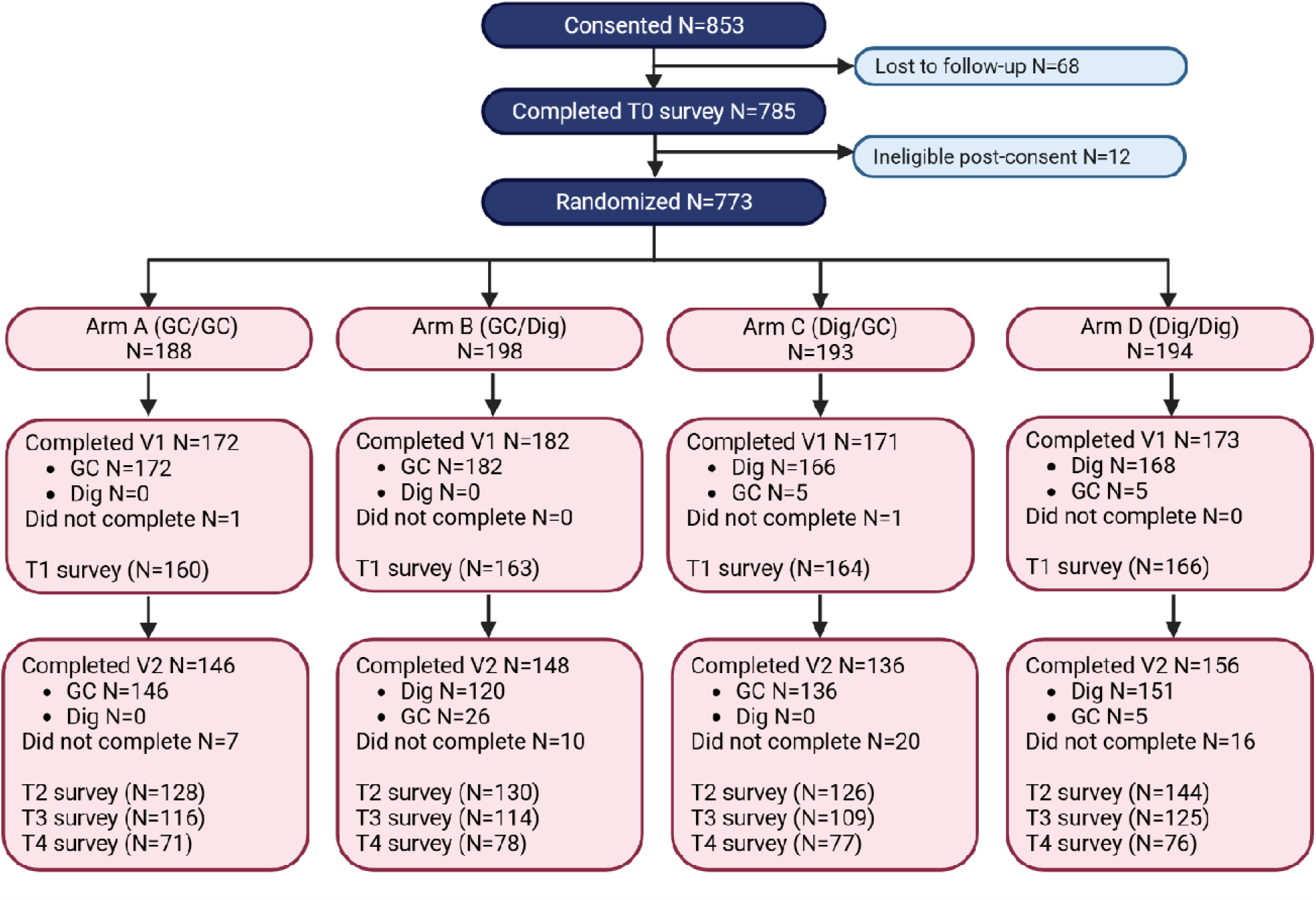
Consolidated Standards of Reporting Trials diagram.

**Table 1:**
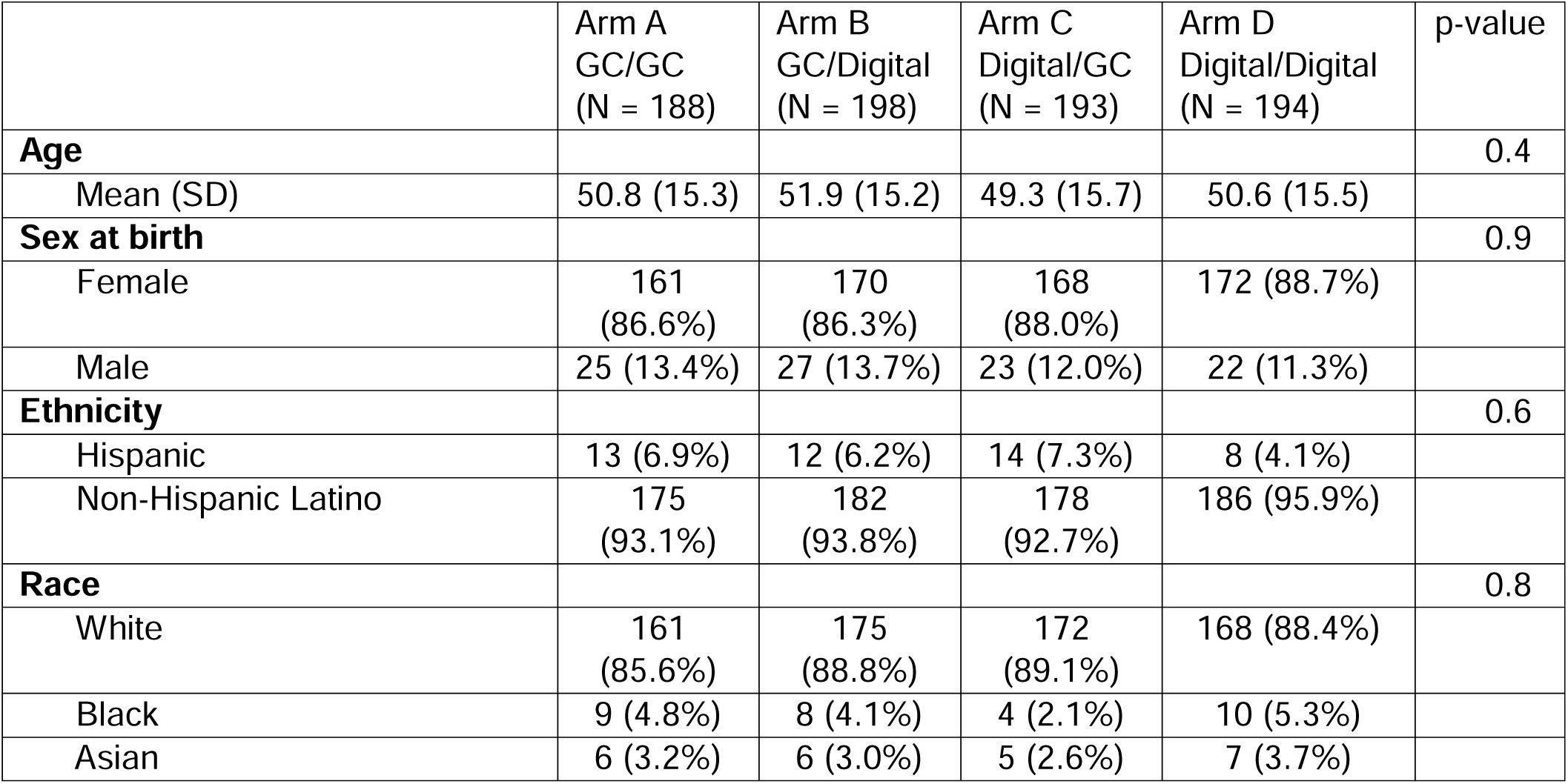
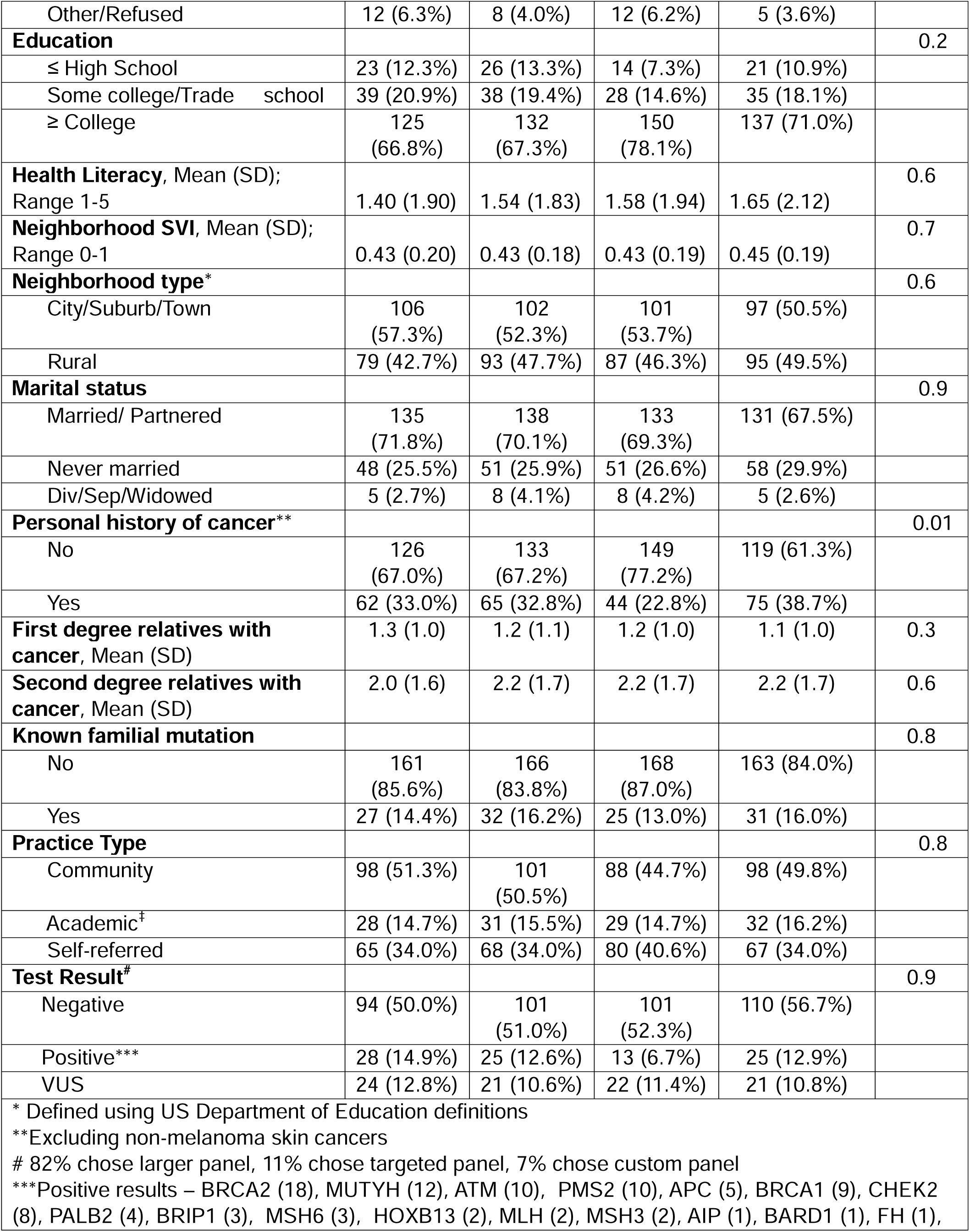
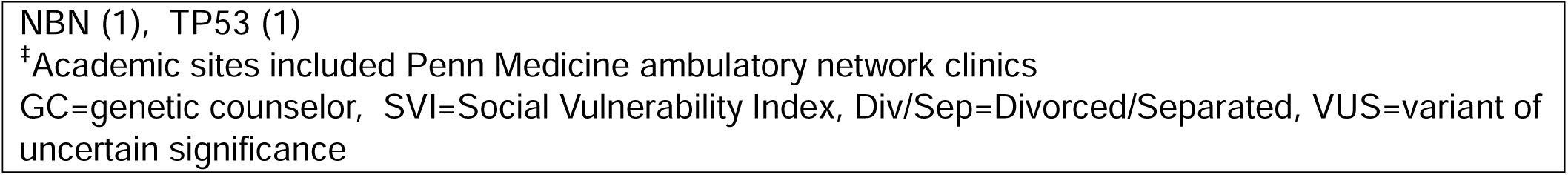
Participant Characteristics at Baseline by Study Arm.

### Non-inferiority analyses and uptake of genetic services

In the primary intention-to-treat (ITT) analyses, we met noninferiority for uptake of testing and anxiety (T2-T0), but results for genetic knowledge (T2-T0) were inconclusive. Standardized effect estimates for each primary outcome are shown in **Figure 2**, and outcome-specific results by study arm are presented in **Table 2**. The secondary endpoints at T2, T3 and T4 exhibited heterogeneous effects. In most cases, arm C demonstrated consistent effects in a beneficial direction, while arms B and D demonstrated effects in an inferior direction.

**Figure 2:**
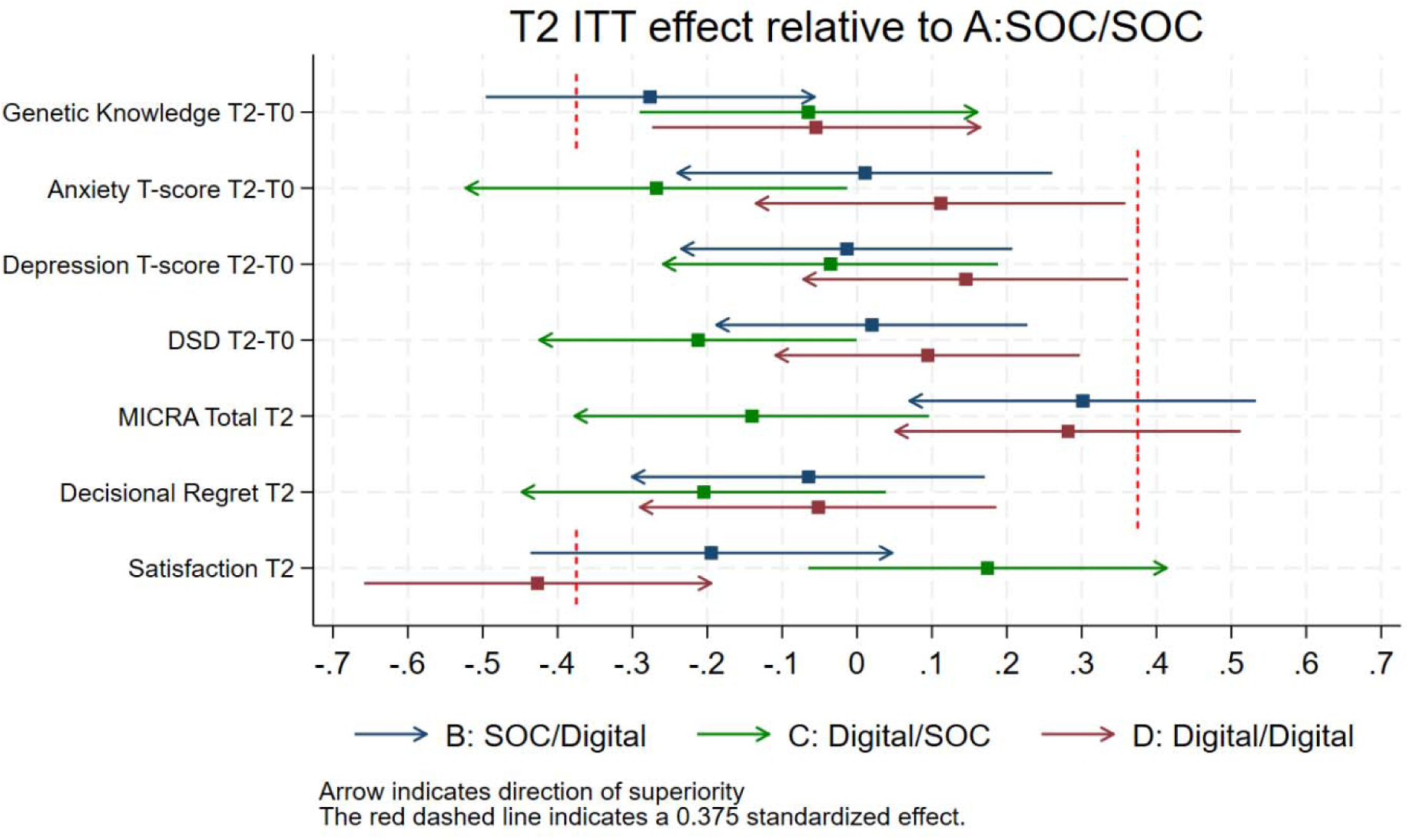
Forest plot depicting prespecified noninferiority margins and confidence intervals by study arm compared to the standard of care arm.

**Table 2:** Mean differences (standard deviation) by arm for the primary and secondary outcomes after visit 1 (T1), visit 2 (T2), 6-months, and 12-months.

| Outcomes, mean (SD) | Arm A<br>GC/GC<br>(n=188) | Arm B<br>GC/Digital<br>(n=198) | Arm C<br>Digital/GC<br>(n=193) | Arm D<br>Digital/Digital<br>(n=194) | P Value<sup>a</sup> |
| --- | --- | --- | --- | --- | --- |
| **Uptake of Visit 1, n, (%)** | 173 (92.0%) | 182 (91.9%) | 170 (88.1%) | 173 (89.2%) | .65 |
| **Uptake of testing n (%)** | 146 (77.7%) | 147 (74.2%) | 136 (70.5%) | 156 (80.4%) | .45 |
| **Uptake of Visit 2, n (%)** | 146 (77.7%) | 146 (73.7%) | 136 (70.5%) | 156 (80.4%) | .19 |
| **Anxiety T-score (T0)** | 52.74 (8.73) | 53.37 (9.39) | 53.82 (8.87) | 53.59 (9.06) | . |
| **Anxiety T-score (T1-T0)** | -3.44 (7.76) | -3.40 (8.85) | -2.33 (8.34) | -2.56 (7.49) | .435 |
| **Anxiety T-score (T2-T0)** | -5.20 (8.88) | -5.18 (10.26) | -7.61 (9.30) | -4.19 (10.23) | **.025** |
| **Anxiety T-score (T3-T0)** | -2.14 (9.17) | -0.64 (8.55) | -1.12 (7.96) | -0.71 (8.91) | .461 |
| **Anxiety T-score (T4-T0)** | -0.30 (8.55) | -0.30 (8.86) | -0.18 (8.59) | -0.15 (9.17) | .999 |
| **Knowledge (T0), range 0-16** | 8.70 (3.55) | 9.58 (3.31) | 9.56 (3.16) | 9.34 (3.54) |  |
| **Knowledge (T1-T0)** | 2.09 (3.17) | 1.61 (2.99) | 2.14 (3.15) | 2.18 (3.24) | .275 |
| **Knowledge (T2-T0)** | 2.34 (3.02) | 1.40 (3.28) | 2.12 (3.22) | 2.15 (3.17) | .069 |
| **Knowledge (T3-T0)** | 1.96 (3.00) | 0.64 (3.13) | 1.58 (3.02) | 1.77 (2.70) | **.001** |
| **Knowledge (T4-T0)** | 1.72 (3.11) | 0.71 (3.38) | 1.39 (3.33) | 1.48 (3.30) | .086 |
| **Depression T-score (T0)** | 48.12 (7.53) | 49.12 (8.23) | 48.40 (8.29) | 48.66 (8.03) |  |
| **Depression T-score (T1-T0)** | -1.79 (6.77) | -2.35 (6.94) | -1.09 (6.67) | -1.26 (6.63) | .270 |
| **Depression T-score (T2-T0)** | -3.15 (7.80) | -3.28 (7.67) | -3.43 (7.06) | -1.98 (7.79) | .344 |
| **Depression T-score (T3-T0)** | 0.93 (7.79) | 0.86 (7.28) | 0.89 (7.64) | 1.31 (7.55) | .955 |
| **Depression T-score (T4-T0)** | 1.67 (7.54) | 1.35 (7.41) | 1.76 (7.65) | 1.93 (7.15) | .934 |
| **Disease specific distress (T0), range 0-40** | 9.89 (9.16) | 9.71 (10.16) | 10.43 (10.02) | 10.73 (9.67) |  |
| **Disease specific distress (T1-T0)** | -2.00 (7.59) | -2.28 (6.79) | -1.53 (7.05) | -1.29 (6.86) | .546 |
| **Disease specific distress (T2-T0)** | -3.16 (8.38) | -2.99 (8.93) | -5.23 (8.63) | -2.23 (8.63) | **.030** |
| **Disease specific distress (T3-T0)** | -2.21 (8.30) | -2.15 (8.50) | -2.51 (9.42) | -2.34 (8.53) | .988 |
| **Disease specific distress (T4-T0)** | -2.34 (8.43) | -1.30 (8.88) | -2.53 (9.59) | -1.86 (8.59) | .733 |
| **Satisfaction with services (T1), range 8-40** | 34.20 (4.09) | 34.19 (4.39) | 32.52 (4.05) | 32.19 (4.26) | **<.001** |
| **Satisfaction with services (T2)** | 34.52 (4.21) | 33.60 (4.60) | 35.31 (4.17) | 32.59 (4.61) | **<.001** |
| **Decisional regret (T2), range 5-25** | 2.54 (4.73) | 2.29 (4.32) | 1.69 (3.57) | 2.33 (3.96) | .388 |
| **Decisional regret (T3)** | 1.93 (3.00) | 2.02 (3.27) | 2.01 (3.51) | 2.40 (3.34) | .636 |
| **Decisional regret (T4)** | 1.66 (1.90) | 1.80 (2.03) | 1.07 (1.40) | 2.28 (2.53) | **<.001** |
| **Responses to testing (MICRA, T2), range 0-95** | 9.19 (9.02) | 12.33 (11.34) | 7.71 (7.82) | 12.15 (12.32) | **<.001** |
| **Responses to testing (MICRA, T3)** | 13.36 (8.74) | 16.28 (11.53) | 12.64 (9.73) | 15.50 (10.16) | **.012** |
| **Responses to testing (MICRA, T4)** | 14.83 (7.26) | 17.20 (9.90) | 14.51 (7.99) | 16.11 (8.73) | .077 |
| <sup>a</sup> p value for the 4 group comparison (superiority, this does not reflect non-inferiority testing) |  |  |  |  |  |

Some participants randomized to a digital visit requested a GC (visit 1-2.9%; visit 2-3.2%). The per-protocol and as-treated analyses showed similar heterogeneity in effects.

Participants who did not complete V1 had higher baseline anxiety than those who did (Promis anxiety T-score 56.2 vs 53.1, p=0.005) and were more likely to not have a usual source of health care such as a primary care provider (88% vs 96%, p=0.005). There were no other sociodemographic or clinical characteristics associated with uptake of genetic services. Participants who did not have genetic testing had higher baseline anxiety and depression scores, reported being more worried about being able to meet normal living expenses, were less confident with being able to handle a financial emergency, were more likely to be recruited from social media, and were less likely to have a personal history of cancer.

### Secondary Between⍰Arm Comparisons

In secondary analyses, we evaluated if pairwise differences between the arms as non-inferiority does not exclude the possibility that some arms perform better than others and could explain inconclusive non-inferiority tests. As shown in **Table 2** (unstandardized effects), decreases in anxiety and disease specific distress were greater in Arm C compared to Arms A, B, and D at T2, but these differences were small and may not be clinically significant and there was no difference at 6- and 12-months. Similarly, participants in Arm C had less negative response to testing (as measured by MICRA) than those in other arms at T2 and 6 months with no difference between arms at 12 months, though MICRA scores were relatively low in all arms. There were no differences in knowledge in the short-term (T0-T1 and T0-T2). At 6 months, knowledge was higher for Arms A, C, and D compared to Arm B, but at 12-months there was no difference between arms. While decisional regret was low overall and was similar between groups at T1-T3, at 12 months, it was higher in Arm D compared to the other arms. While satisfaction was slightly higher in arms with GC-delivered education at T1 and T2, the absolute differences were small and likely not clinically meaningful given the consistently high scores observed across all arms. Findings were similar in as-treated analyses.

### Secondary outcomes by test result

Among participants who received a negative result (**Supplemental Table 1**), genetic knowledge gains were similar across arms overall, with a transient decrease in Arm B at T3 that attenuated by T4. All arms demonstrated decreases in anxiety and disease specific distress relative to baseline, but Arm C showed the greatest reduction at T2. Satisfaction was slightly higher with GC visits compared to digital visits, although the differences were small, satisfaction was high in all arms and these differences are likely not clinically significant. MICRA scores were low overall, and while MICRA scores were higher for participants in Arm B compared to the other arms, the absolute differences were small, and not likely clinically significant. Decisional regret was generally similar across arms, with a small increase in Arm D at T4 that may not be clinically significant.

Among participants who received a VUS result (**Supplemental Table 1**), knowledge gains were comparable across arms, with only a transient reduction in Arm B at T3 that resolved by T4. Short-term reductions in anxiety and disease-specific distress were similar in Arms A and C but smaller in Arms B and D; however, these differences were not sustained at later follow-up. At T2, MICRA scores were low overall and lower for Arms A and C than the digital disclosure arms (B and D). The observed MICRA differences were approximately 0.5 standard deviation units for VUS results at T2, indicating moderate standardized effects, but the magnitude of differences were attenuated at T3 and T4. Decisional regret was generally similar between arms, with only a small increase in Arms B and D at T4.

Among participants who received a positive result (**Supplemental Table 1**), overall, Arm C produced outcomes most similar to Arm A, while Arms B and D showed some evidence of less favorable patient-reported outcomes. Genetic knowledge gains were comparable across arms, with only transient reductions in Arms B and C at T3. There was a qualitative difference at T2 for anxiety and disease-specific distress outcomes, with arms A and C showing reductions, but arms B and D having increases; these differences were not sustained over time. Satisfaction was slightly higher for GC visits. Lowest MICRA scores were observed in Arms A and C and the difference between arms A and C compared to Arms B and D T2 are larger than for other findings, and the absolute scores are high enough to be potentially clinically significant. The observed differences are greater than one standard deviation units, indicating large standardized effects. Differences between decisional regret were unlikely to be clinically significant among arms.

### Moderators of patient reported outcomes and testing

There were no significant moderators of PROs for pre-test counseling (visit 1, T0-T1). For disclosure visits (T1-T2), test result was a moderator of MICRA. Those with positive results had higher MICRA scores at T2 with digital disclosure (24.8-Arm B and 26.4-Arm D) compared with GC disclosure (15.1-Arm A and 17.4-Arm C, **Supplemental Table 1**). Those with VUS results had slightly higher MICRA scores at T2 with digital disclosure, but these are less likely clinically significant (Arm B-14.9 and Arm D-18.2 vs Arm A-9.4 and Arm C 9.6). Those with lower health literacy had higher test related uncertainty in the fully digital arm.

### Genetic counselor time and intervention use

GCs spent the most time on care for participants in Arm A and the least time for participants in Arm C, as shown in **Figure 3**. Specific GC activities and time spent are available in **Supplemental Table 2**.

**Figure 3:**
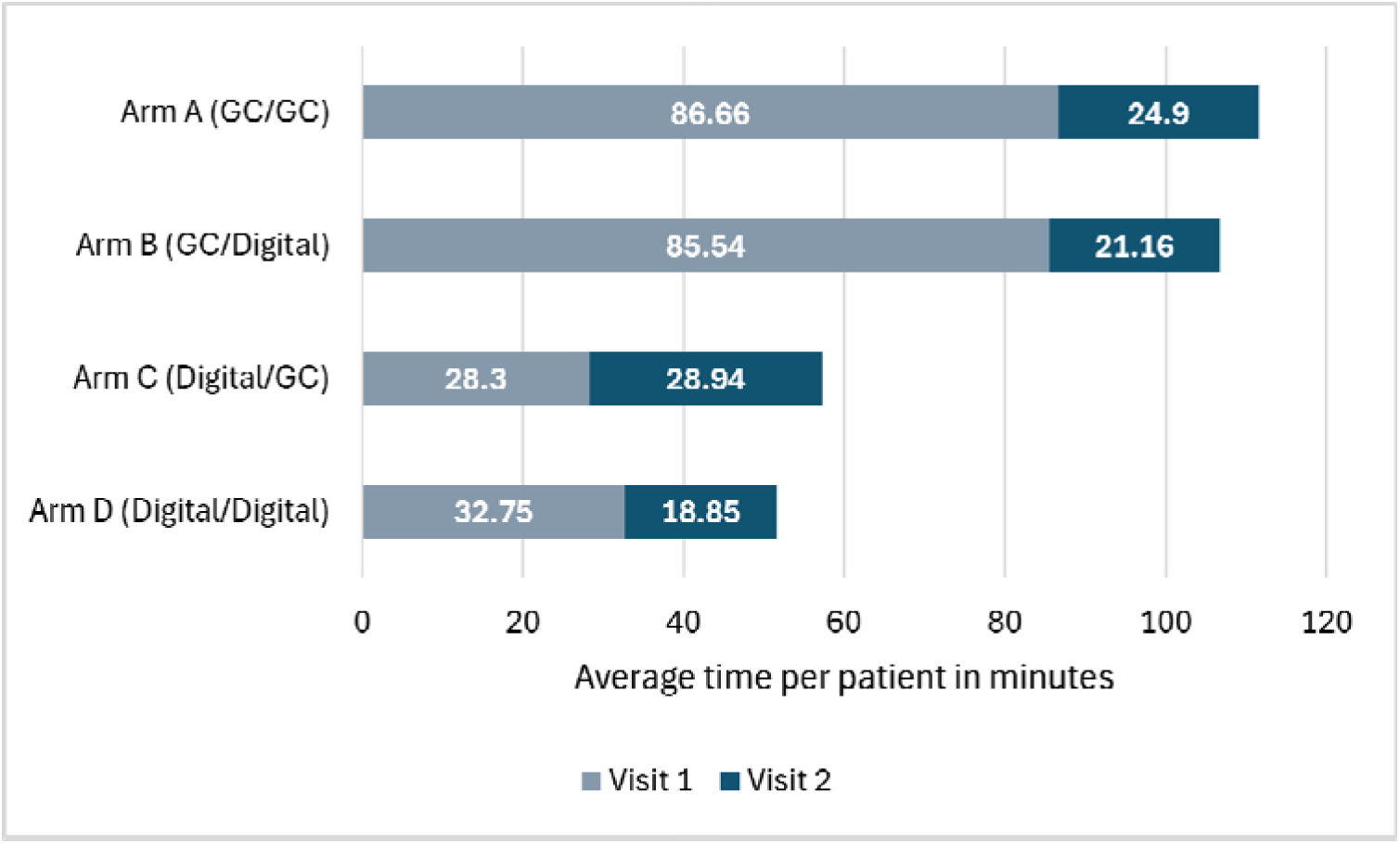
Genetic counselor time per patient by randomized arm and visit

## DISCUSSION

This randomized non-inferiority trial evaluated a patient-centered digital alternative to GC for pre-test counseling and/or results disclosure in real-world patients who qualify for germline cancer genetic testing. We found that across a diverse sample of participants, the eREACH intervention was effective for pre-test counseling, but inconclusive for digital disclosure of results. Critically, Arm C (digital pre-test education followed by GC-delivered disclosure) demonstrated similar outcomes to two visits with a GC over all other arms in reducing anxiety, disease-specific distress, and negative responses to testing, and even among participants receiving a positive or VUS result. In contrast, among participants who received a negative result, outcomes were broadly similar across all arms, with only small and likely clinically insignificant differences observed, suggesting that digital disclosure may be an appropriate and acceptable model for this population. Taken together, these findings support the feasibility of integrating digital tools into genetic services workflows, particularly for pre-test education and counseling, while underscoring the potential continued importance of GC involvement in the disclosure of clinically actionable cancer genetic results.

Our findings add context to other randomized studies evaluating alternative delivery models in clinical genetics. eREACH1 established the evidence for the digital intervention tested in this trial by demonstrating non-inferiority of digital delivery of pre-test counseling or post-test result disclosure compared to two visits with a GC among patients with certain metastatic cancers.[15] eREACH2 extends the eREACH1 findings substantially by expanding beyond the metastatic cancer setting to a broad real-world population of patients with and without cancer meeting NCCN and ASCO guidelines for genetic testing. While prior randomized studies largely focused on whether digital approaches could replace portions of the pre-test pathway, eREACH2 evaluates whether digital models can be extended to return of results and whether outcomes differ based on the genetic test result itself. The MAGENTA study found that an educational video and omitting mandatory post-test counseling for an unaffected, female-only population without a PV was non-inferior to GC visits in terms of distress, but did not evaluate patient responses to testing as measured by MICRA — an important affective outcomes assessed in eREACH2.[32] Similarly, the BRIDGE study found that a pre-test chatbot was equivalent to GC pre-test counseling for uptake of cancer genetic services, but did not evaluate the impact of digital disclosure on post-test patient-reported outcomes.[33] BRCA-DIRECT demonstrated non-inferiority of digital delivery of pre-test information for knowledge and anxiety compared to a telephone GC consultation, findings that generally align with our results. However, all participants with positive results received GC disclosure, the intervention focused pre-test information delivery, the study was restricted to female breast cancer patients undergoing BRCA1/2 and PALB2 testing within a single healthcare system, and it did not include MICRA, where we observed important differences. Together, these studies established that digital approaches can effectively support pre-test education and facilitate testing uptake. Importantly, eREACH2 is the first large randomized study to evaluate outcomes of digital disclosure among individuals receiving PV and VUS results, allowing assessment of patient responses to testing and other post-disclosure outcomes. Our study extends prior work by including men, a wide range of patients meeting guidelines for cancer genetic testing, including participants with and without a personal history of cancer and individuals with a known familial mutation. We also evaluated outcomes not previously examined in many trials, including responses to testing (MICRA), patient preferences for digital versus GC delivery, and provider time requirements. Furthermore, we utilized real-world testing practices in which insurance was billed and patients were responsible for any out-of-pocket costs, recruited nationally and included patients in community practices and rural settings.

One major strength of eREACH 2 was the study size and ability to evaluate outcomes by genetic test results in secondary analyses, the first large study of a digital delivery intervention to do so. We found that some patient outcomes with digital disclosure may vary by genetic test result, providing important nuances for clinical implementation. Among participants who received a negative result, outcomes were broadly comparable across all four arms, with no clinically significant differences in knowledge, anxiety, distress, or decisional regret. These findings support the safety and appropriateness of fully digital delivery (Arm D) for patients ultimately receiving a negative result, representing the majority of tested participants (69.4%), and suggesting that the resource intensity of GC involvement may not be necessary for this group. Among participants who received a VUS, results were similarly reassuring, with comparable knowledge gains and only transient, non-sustained differences in anxiety and distress in Arms B and D. While the differences in digital disclosure were modest and could be considered clinically significant, the absolute scores were still relatively low. These findings suggest that digital delivery is a reasonable and efficient option for patients receiving VUS results, though clinicians may consider patient-specific factors — such as elevated baseline anxiety or limited health literacy — when determining whether additional GC support is warranted. Patients with a positive result require greatest caution in implementation decisions. Arm C consistently produced outcomes most similar to Arm A (GC/GC), suggesting that GC disclosure may provide some benefit for this group. Although increases in MICRA scores with digital disclosure were transient, the short term increases in negative responses to testing could be considered clinically significant. Of note, these were exploratory analyses and not powered for definitive subgroup comparisons. Thus, clinicians may want to weigh the risks and benefits of digital disclosure for individuals with VUS and positive results. When possible, maintaining GC disclosure may be prudent. But, for selected patients facing substantial barriers to genetics services this patient-centered digital intervention could be considered for willing patients.

Very few participants randomized to digital visits requested a GC (2.9% at visit 1; 3.2% at visit 2) suggesting that the digital intervention was acceptable to participants. These rates are substantially lower than those observed in the eREACH1 study, in which 11.1% of participants requested a GC for visit 1 and 14.5% for visit 2.[15] This difference may be due to differences in the study population and clinical context across the two trials. GC time was substantially reduced when digital interventions replaced one or both GC visits, with the least provider time required in Arm C, the arm that also produced the most consistently favorable patient outcomes. While digital delivery models have the potential to improve access to genetic services without sacrificing patient outcomes, these data highlight the continued need for and value of GC involvement in these models.[18]

The time savings of the digital intervention can increase GC capacity, allowing GCs to redirect their expertise toward higher-complexity cases, positive result disclosures, and other patients who require individualized support. GCs would be able to see more patients, reduce appointment wait times, thereby improving patient access to timely genetic services. Practically, this model would require significant changes to reimbursement policies, allowing for digital genetic services or asynchronous patient engagement as reimbursable components of genetic counseling care. Payers and health systems will need to collaborate to develop reimbursement frameworks that recognize digital delivery models as an evidence-based approach to care that maintains patient outcomes while expanding access. This model also requires a shift in how GCs practice and comfort with this new paradigm may vary. We note that GCs remain indispensable in this model, shifting only in the nature of their role to maximize their value. Further work is needed to understand concerns about this framework in the GC community.

We acknowledge some limitations. While this was a prospective, randomized, real-world trial with extensive PROs, and participants recruited from 46 states with 49% enrollment from community setting, and we were able to examine outcomes by test results, these were secondary analyses and the study was not powered on these result specific endpoints. Further analysis to determine post-test health behavior and perceived risk assessment may provide additional insight into the use of the fully digital model for those with negative results. GC time data was collected for a small subset of participants which may limit the precision of these estimates. The proposed model in the eREACH studies requires changes in how GCs practice and provider acceptability is unknown. Novel reimbursement models for “virtual care” would also be needed for broad dissemination of these tools.

In conclusion, this randomized non-inferiority study demonstrates that the eREACH patient-centered digital intervention is largely non-inferior to traditional GC-led care for key outcomes including anxiety and uptake of genetic services among patients meeting national guidelines for germline genetic testing. For individuals receiving negative results, fully digital delivery appears to be an acceptable alternative to GC care. For those receiving a positive result, however, the data suggest that GC involvement in the disclosure process may remain important to support optimal short-term test-related distress. A hybrid model that leverages digital tools for pre-test education while preserving GC-led disclosure for actionable results may represent the most pragmatic and scalable approach — one that extends the reach of genetic services without compromising care quality for those who may need it most.

## Data Availability

All data produced in the present study are available upon reasonable request to the authors

## Abbreviations

eREACH: A Randomized Study of an eHealth Delivery Alternative for Cancer Genetic Testing for Hereditary Predisposition in Metastatic Breast, Ovarian, Prostate, and Pancreatic Cancer Patients
GC: genetic counselor
HIPAA: Health Insurance Portability and Accountability Act
MAGENTA: Making Genetic Testing Accessible
PARP: poly (adenosine diphosphate ribose) polymerase
REDCap: Research Electronic Data Capture
SPIRIT: Standard Protocol Items Recommendations for Interventional Trials
TARGET: Technology-Enhanced Acceleration of Germline Evaluation for Therapy

## Acknowledgements

KTL’s time was supported by the National Cancer Institute of the National Institutes of Health under award K08CA279076. Generative artificial intelligence was used to edit the manuscript. Figure 1 was created using BioRender.

---

**Supplemental Figure 1.**
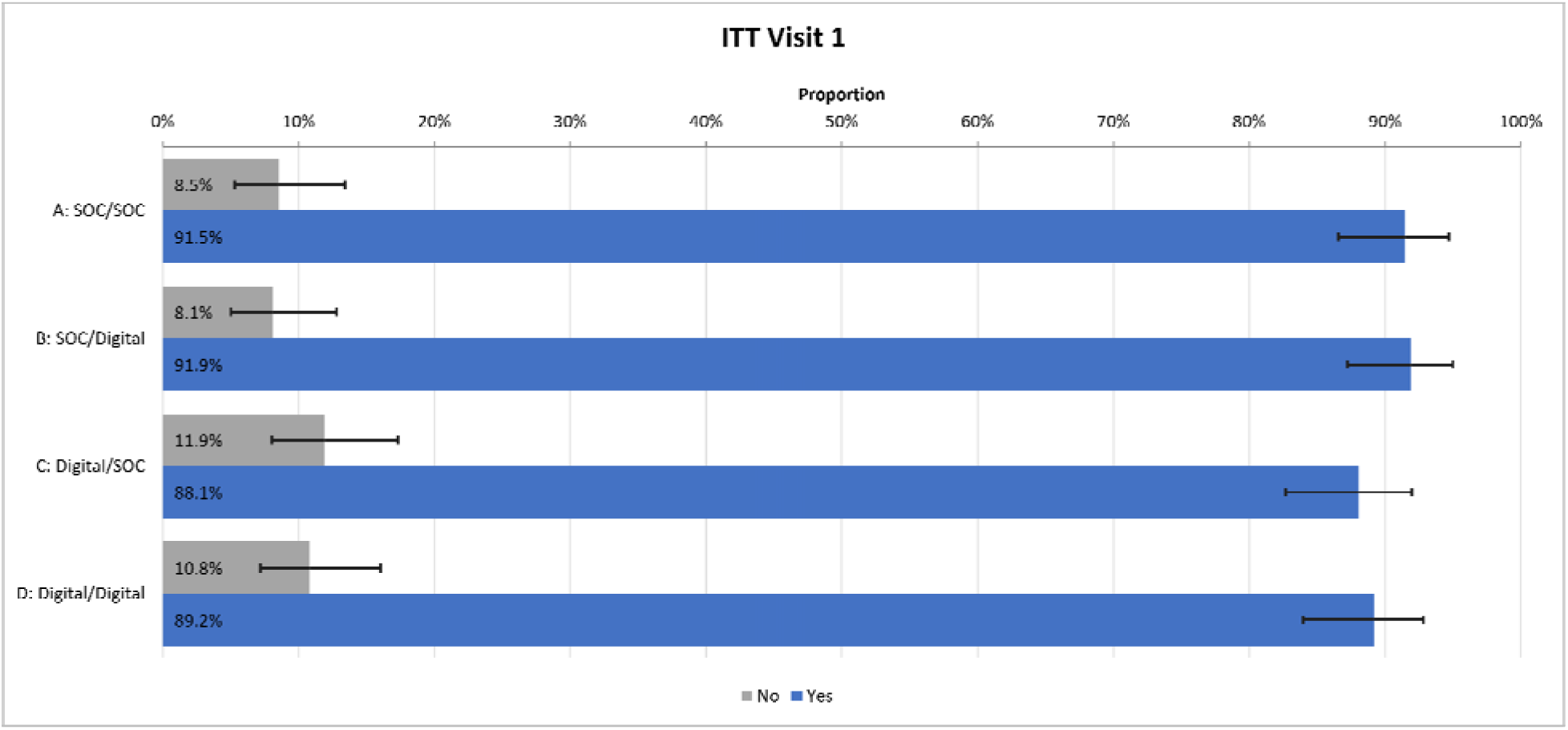
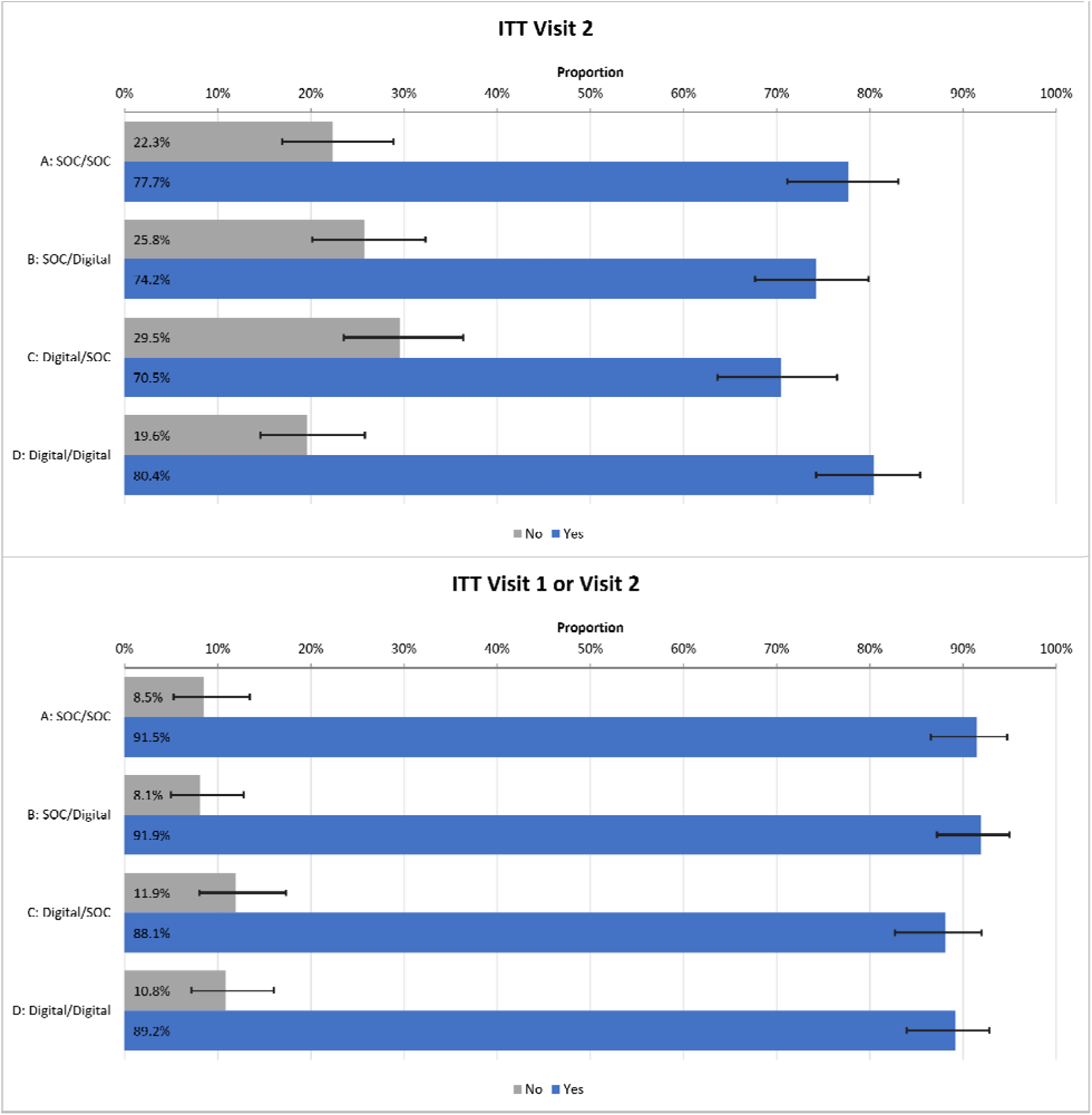
Non-inferiority of uptake of genetic services

**Supplemental Table 1:**
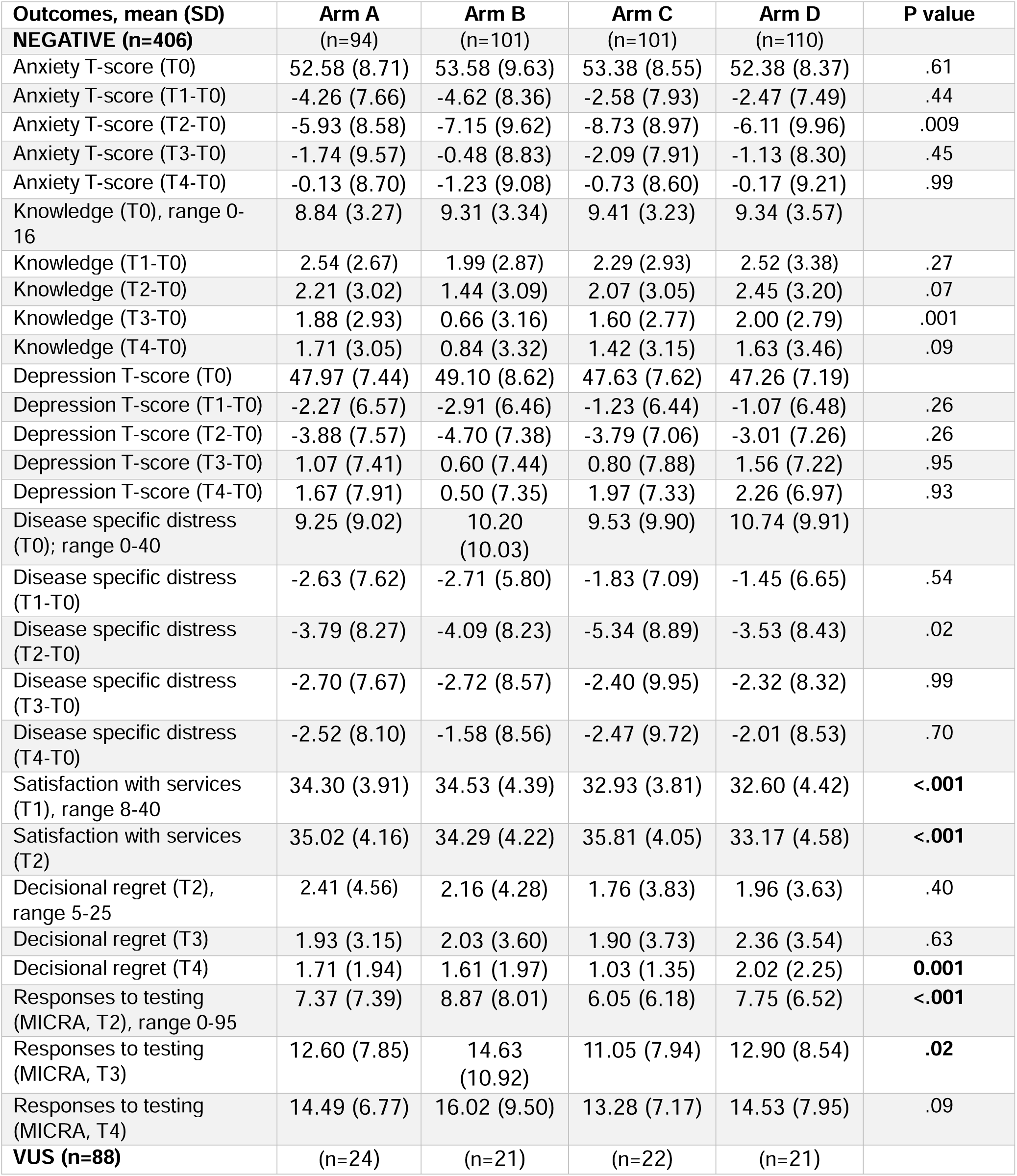
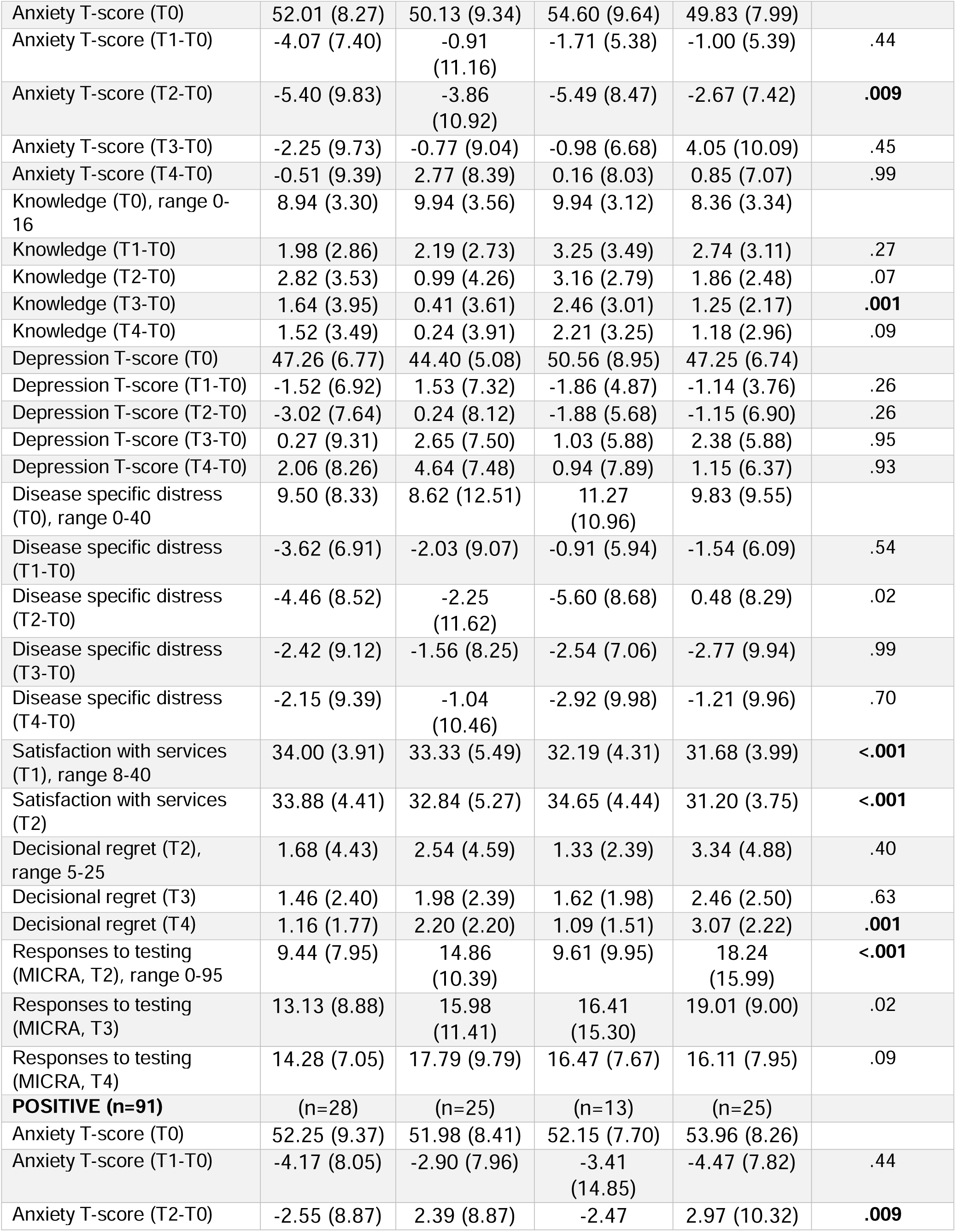
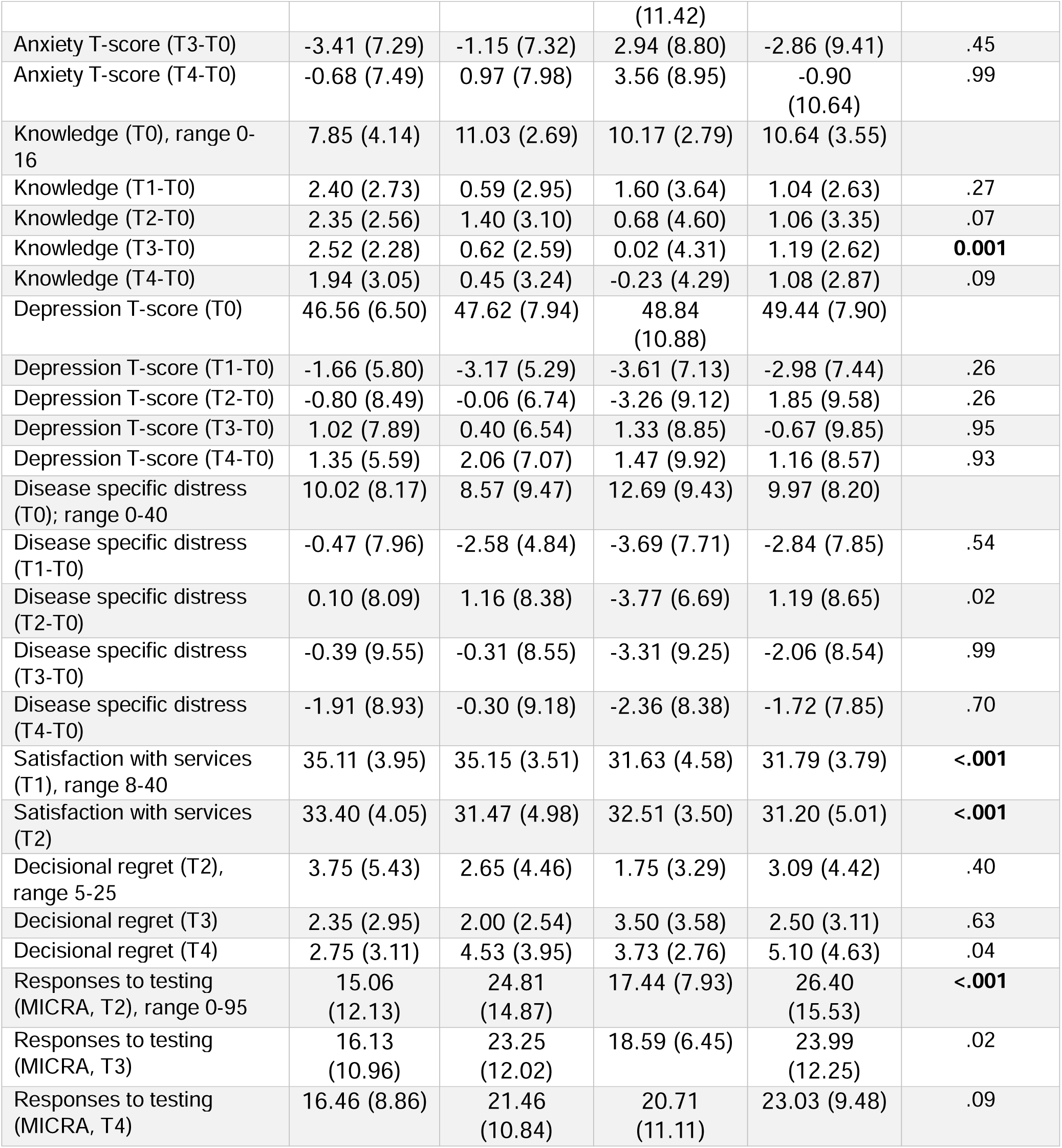
Mean differences by arm for patient reported outcomes by test result.

**Supplemental Table 2:** GC time and effort by visit.

| Visit 1 (average time in minutes) |  |  |  |  |  |
| --- | --- | --- | --- | --- | --- |
| ACTIVITY | ARM A<br>N=35 | ARM B<br>N=37 | ARM C<br>N=27 | ARM D<br>N=24 | Total time by<br>activity |
| Chart Review | 12.91 (6-25) | 12.57 (4-20) | 7.78 (0-20) | 8.92 (0-15) | 10.55 (0-25) |
| Session Time (GC) | 49.91 (25-74) | 47.59 (20-82) | NA | NA | 48.75 (20-82) |
| Communication with Patient After Session (SOC) | 1.37 (0-15) | 2.03 (0-30) | none | none | 1.70 (0-30) |
| Communication with Patient After Session (WEB) | NA | NA | 5.67 (0-45) | 6.42 (0-35) | 6.05 (0-45) |
| Test Order Facilitation+ | 9.11 (0-30) | 10.32 (5-25) | 7.41 (0-15) | 7.25 (0-15) | 8.52 (0-30) |
| Insurance Navigation (Company) | 0.57 (0-15) | 0.27 (0-10) | 1.85 (0-15) | 3.04 (0-20) | 1.43 (0-20) |
| Insurance Navigation (Patient) | 0.14 (0-5) | 0.78 (0-15) | none | 2.29 (0-30) | 0.80 (0-30) |
| Chart Note Documentation+ | 11.60 (0-25) | 11.14 (0-40) | 4.67 (0-17) | 4.63 (0-13) | 8.01 (0-40) |
| Other | 1.03 (0-26) | 0.84 (0-10) | 0.93 (0-25) | 0.21 (0-5) | 0.75 (0-26) |
| V1 total time | 86.66 (56-147) | 85.54 (45-148) | 28.30 (15-92) | 32.75 (10-65) | 58.31 (10-148) |
| Visit 2 (average time in minutes) |  |  |  |  |  |
| ACTIVITY | ARM A<br>N=29 | ARM B<br>N=32 | ARM C<br>N=19 | ARM D<br>N=20 | Total time by<br>activity |
| Result Interpretation+ | 1.71 (0-10) | 2.65 (0-20) | 3.31 (0-15) | 2.35 (0-10) | 2.51 (0-20) |
| Approval of Web Result+ | NA | 0.69 (0-5) | NA | 0 | 0.34 (0-5) |
| Chart Review+ | 1.28 (0-13) | 1.16 (0-10) | 2.06 (0-8) | 2.00 (0-10) | 1.63 (0-13) |
| Session Time (GC session*) | 9.59 (4-28) | 2.77 (0-30) | 12.00 (4-30) | 3.60 (0-40) | 6.99 (0-40) |
| Communication with Patient (Post Disclosure Session) | 0.69 (0-10) | 1.71 (0-20) | None | 1.00 (0-10) | 0.85 (0-20) |
| Chart Note Documentation | 11.24 (5-25) | 11.74 (5-35) | 11.56 (5-25) | 9.90 (5-20) | 11.11 (5-35) |
| Post Disclosure Communication w/ Provider | 0 | 0 | 0 | 0 | 0 |
| Other | 0.10 (0-3) | 1.13 (0-30) | 0 | 0 | 0.31 (0-30) |
| V2 total time | 24.90 (10-73) | 21.16 (5-70) | 28.94 (9-52) | 18.85 (5-65) | 23.46 (5-73) |
| **Combined total time** | **111.97 (68-174)** | **104.50 (55-202)** | **45.44 (15-129)** | **48.46 (15-112)** | **77.59 (15-202)** |

^Not all individuals tested
+Not all GCs included this as part of time tracking
\*Includes individuals who opted out of digital disclosure (Arms B and D)

